# Transmissibility of coronavirus disease 2019 (COVID-19) in Chinese cities with different transmission dynamics of imported cases

**DOI:** 10.1101/2020.03.15.20036541

**Authors:** Ka Chun Chong, Wei Cheng, Shi Zhao, Feng Ling, Kirran N. Mohammad, Maggie Haitian Wang, Benny Chung Ying Zee, Lei Wei, Xi Xiong, Hengyan Liu, Jingxuan Wang, Enfu Chen

## Abstract

**Background:** Monitoring the time-varying reproduction number (*R*_*t*_) of the disease is useful in determining whether there is sustained transmission in a population. In this study, we examined *R*_*t*_ of COVID-19 and compared its transmissibility between different intervention periods in Hangzhou and Shenzhen.

**Methods:** Daily aggregated counts of confirmed imported and local cases between January 1, 2020 and March 13, 2020 were analysed. A likelihood function was constructed to estimate *R*_*t*_, accounting for imported cases.

**Results:** Although Hangzhou had fewer number of cases than Shenzhen, Shenzhen had higher proportion of imported cases than Hangzhou (83% vs 29%). Since the epidemic of COVID-19 in Shenzhen was dominated by imported cases, *R*_*t*_ was kept below unity through time. On the contrary, *R*_*t*_ was greater than unity in Hangzhou from 16 January to 7 February due to the surge in local cases. Credits to the Wuhan lockdown and outbreak response measures following the local lockdown, *R*_*t*_ decreased steadily and dropped below unity in mid-February.

**Conclusion:** The lockdown measures and local outbreak responses helped reduce the potential of local transmission in Hangzhou and Shenzhen. Meanwhile, cities with similar epidemic trend could have different transmission dynamics given the variation in imported cases.

## Backgrounds

Coronaviruses are a diverse group of enveloped, positive-sense, single-stranded RNA viruses that belongs to the family Coronaviridae, order Nidovirales [1]. The diseases caused by these viruses are zoonotic in nature, which can be transmitted between animals and human [2]. In human, coronaviruses mainly cause respiratory tract infections with severe acute respiratory syndrome (SARS) and Middle East respiratory syndrome (MERS) being two notable examples. In December 2019, a novel strain of coronavirus that has not been previously identified in human emerged and caused an outbreak in Wuhan, Hubei province, China [3]. The disease responsible for the outbreak has been officially named by the World Health Organization as coronavirus disease 2019 (COVID-19). Common clinical manifestation of COVID-19 includes fever, fatigue, dry cough, shortness of breath and muscle ache [4,5]. In severe cases, COVID-19 may progress to pneumonia or even death. Symptoms such as headache, dizziness, sputum production, haemoptysis, diarrhoea, nausea and vomiting are less common but occasionally reported. According to the Centers for Disease Control and Prevention, COVID-19 is mainly spread from person-to-person via respiratory droplets, but may also be spread from contact with infected surfaces or objects [6]. At the moment, there is no vaccine nor specific antiviral treatment approved for COVID-19.

The current global scale health crisis triggered by COVID-19 began on 31 December 2019, the day the World Health Organization was notified of several severe and unusual cases of pneumonia in Wuhan. Despite the suspected source of transmission (Huanan Seafood Wholesale Market) was shut down the next day, number of cases continued to expand at an alarming rate [7]. Health experts soon identified the cause of the disease as being a novel coronavirus that can be spread via airborne droplets and announced the discovery on 7 January 2020. Two days later, the first related death was recorded in Wuhan; meanwhile, the disease spread outside Hubei as people travel around and outside the country, especially during “Chunyun”, a 40-day period when Chinese people going home to celebrate the Lunar New Year holidays. On 13 January 2020, Thailand reported the first lab-confirmed case of COVID-19 outside mainland China and health authorities of 19 other countries confirmed cases over the following days [8]. By 31 January 2020, a total of 9,826 cases of COVID-19 were confirmed around the globe and the figure surpassed 10,000 the next day.

To halt the spread of the virus, the Chinese government imposed a complete lockdown in Wuhan and other cities in Hubei province on 23 January 2020 aiming to quarantine the epicenter of the COVID-19 outbreak [9]. All public transportation, including buses, railways, flights, and ferry services, was suspended and residents were forbidden from leaving the city without permission from the authorities. Later, the government tightened its quarantine measures and imposed a mandatory 14-day quarantine on people returning to the capital from holidays. Meanwhile, a number of countries have imposed restrictions on entry by travellers from China in response to the epidemic. Several early investigations have shown that the shutdown of cities largely reduced the spread of infections to other cities in China and other countries around the world [10-13]. Time-varying reproduction number (*R*_*t*_) is defined as the average number of secondary cases infected by a primary case at time *t. R*_*t*_ is a common measure for monitoring the disease evolution in a population especially when some control measures have been carried out to mitigate an epidemic. *R*_*t*_ below unity indicates a single case is unable to produce more than one secondary case on average and the disease can unlikely be sustained in a population over time. In this case, the outbreak can be regarded as under control at time *t* [14]. In this study, we examined the transmission dynamics of COVID-19 during the first wave of epidemic and compared its transmissibility between different intervention periods in Hangzhou and Shenzhen, two Chinese cities with different transmission dynamics of imported cases.

## Methods

### Setting

Hangzhou and Shenzhen are two major Chinese cities in terms of economy, population and transportation. Hangzhou is the capital city of Zhejiang Province and one of the most populous cities in East China. In 2018, the city’s resident population was 9.8 million with an estimation of 4 million floating population [15]. With 16,424 km length of highways, well-developed railway transportation and 286 civil aviation routes between 160 domestic cities, the passenger traffic of Hangzhou was over 20 million in 2018 [16]. Similar to Hangzhou, Shenzhen is also a new-growing city characterized by high-technology industry, large population of “migrant workers” and well-developed transportation network. Located in Guangdong Province, it covers an area of around 2,000 km^2^ and is the most populous city in South China. In 2018, the number of people living in Shenzhen was estimated to be 25.7 million, among whom less than half were long-term residents [17].

Wuhan, the source city of COVID-19, was lockdown on 23 January 2020. On the same day, Hangzhou and Shenzhen declared first-level public health emergency in response to the outbreak. Measures including promotion of personal hygiene, cessation of crowd gathering activities, 7-day home quarantine of people from Hubei, restrictions of public transports and mandatory quarantine for close contacts were adopted. On 4 February 2020, a lockdown was imposed in Hangzhou and only one person per household is allowed to leave their home every two days. Public gatherings such as funerals and weddings were banned; all public venues and work places were closed. On 7 February 2020, all living communities in Shenzhen were requested to lockdown in order to contain the local outbreak [18].

### Data collection

We obtained the daily aggregated data of laboratory confirmed COVID-19 cases in Hangzhou and Shenzhen between January 1, 2020 and March 13, 2020 from the National Health Commission and other official websites [18-20]. The date of illness onset (defined by the first appearance of COVID-19 related symptoms) was used to construct the epidemic curves. Cases with travel history to other provinces (with ≥1 case reported) 14 days prior to symptoms onset are defined as import cases.

### Estimation of reproduction numbers

Since the COVID-19 was seeded by the imported cases in many cities outside Hubei, the estimation equation from Thompson et al. [21], an alternative to Wallinga and Teunis [22], was used, accounting for the risk of imported cases [23].

The total number of reported cases (*I(t)*) on day *t* (*t* = 0, 1, 2, 3…) is the sum of imported cases (*h*_*I*_*(t)*) and local cases (*h*_*L*_*(t)*). The total number of infections on day *t* can be computed as

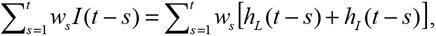

given *w*_*s*_ is a discretized probability distribution function of the serial interval of COVID-19. The expected number of local cases, *E[h*_*L*_*(t)]*, is

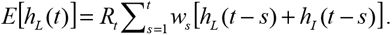

Assuming the number of local infections follows a Poisson distribution, a likelihood function, *L(*·), can be formed as follows

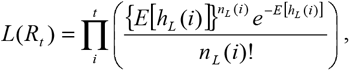

where *n*_*L*_*(t)* is the observed numbers of local cases on day *t*.

We used Markov chain Monte Carlo (MCMC) method to estimate the *R*_*t*_ series based on the observed epidemic curves. Using an early estimate of serial interval [13], we assumed the length of the serial interval follows a lognormal distribution with a mean of 4.4 days and a standard deviation (SD) of 3 days. The sensitivity of results to shorter serial interval of 2.0 days (SD: 2.8) [24] and longer serial interval of 7.5 days (SD: 3.4) [25] was tested. A random walk Metropolis algorithm was used to obtain the posterior distributions of *R*_*t*_. Step sizes were selected to obtain acceptance proportions of 20 to 40%. Twenty thousand MCMC iterations were used as the burn-in period and subsequent 100,000 iterations were used to obtain the estimates. The median and 95% credible intervals (CIs) were calculated to summarize the estimates. The analysis in this study was carried out using software R (version 3.6.3).

## Results

The temporal distribution of cases and *R*_*t*_s in Shenzhen and Hangzhou were shown in Figure 1 and Figure 2 respectively. Shenzhen had the first imported case with illness onset on 1 January, whereas Hangzhou had the first imported case with illness onset on 13 January. From 1 January to 13 March, there were a total of 169 and 417 confirmed cases in Hangzhou and Shenzhen respectively. Both cities had their peak incidence of cases in the week between 22 and 29 January and the epidemics died out in mid-February. In general, although Hangzhou had a fewer number of cases than Shenzhen, Hangzhou had a lower percentage of imported cases than Shenzhen (29% vs 83%). Due to the epidemic in Shenzhen was dominated by imported cases, local *R*_*t*_ was kept below unity through time. In the early phase of the epidemic, *R*_*t*_ was maintained to be around 0.2 to 0.4, indicating a low risk of local transmission despite the rapid increase in daily number of cases before the lockdown of Wuhan (i.e. on 23 January). After the lockdown of Wuhan and the initialization of measures in response to the outbreak, local transmission was well-controlled as indicated by a low estimated value of *R*_*t*_ (i.e. < 0.2). In contrast, *R*_*t*_ was larger than unity in Hangzhou from 16 January to 7 February. Before the lockdown of Wuhan, *R*_*t*_ obtained ranged from 1.9 to 3.8, indicating a high risk of local transmission. Credits to the Wuhan lockdown and outbreak response measures following the local lockdown, *R*_*t*_ dropped steadily and was 1.14 (95% CI: 1.04 to 1.25) on 4 February. After that, *R*_*t*_ kept decreasing slightly and dropped below unity on 10 February, indicating the outbreak was controlled in Hangzhou.

**Figure 1.**
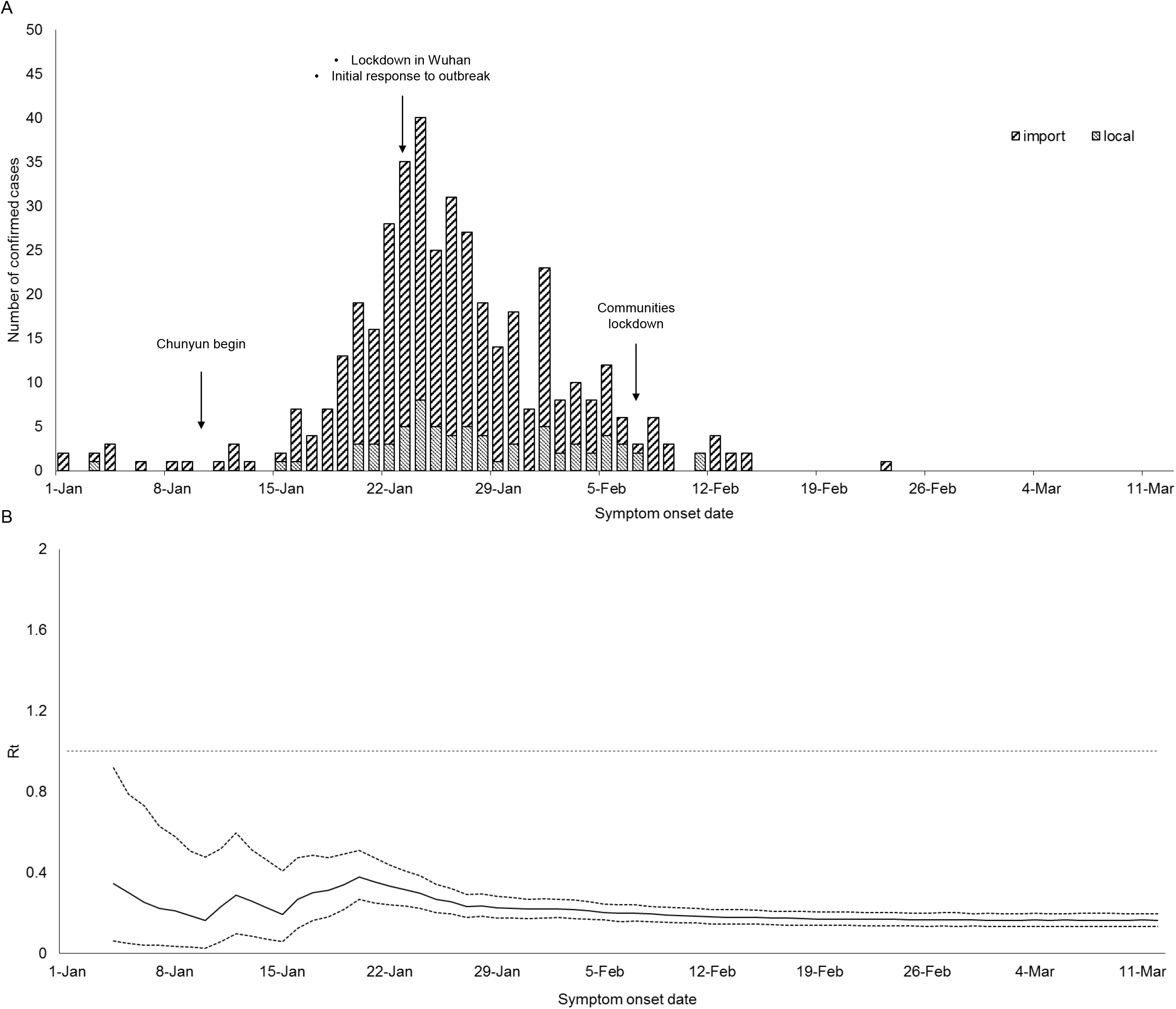
(A) Epidemic trend and (B) Estimate (solid line) and 95% credible intervals (dotted lines) of the time-varying reproduction number (*R*_*t*_) in Hangzhou

**Figure 2.**
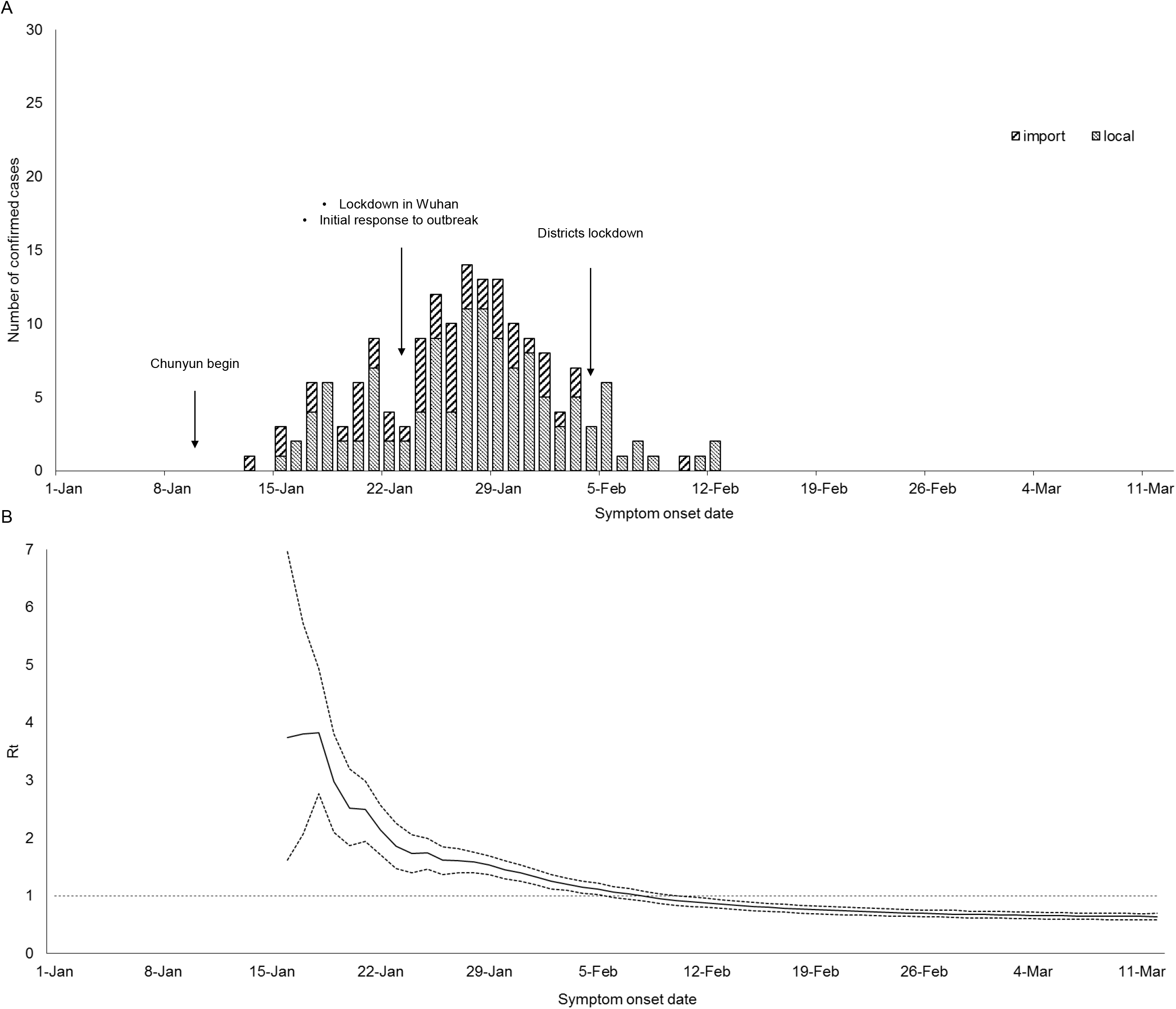
(A) Epidemic trend and (B) Estimate (solid line) and 95% credible intervals (dotted lines) of the time-varying reproduction number (*R*_*t*_) in Shenzhen

Sensitivity analysis was used to assess the robustness of estimated *R*_*t*_ to different serial interval durations (Figure 3). While a variation in the length of serial interval did not affect the estimated *R*_*t*_s in Shenzhen overall, a moderately increased *R*_*t*_ was observed for Hangzhou during the first week when a longer serial interval was assumed.

**Figure 3.**
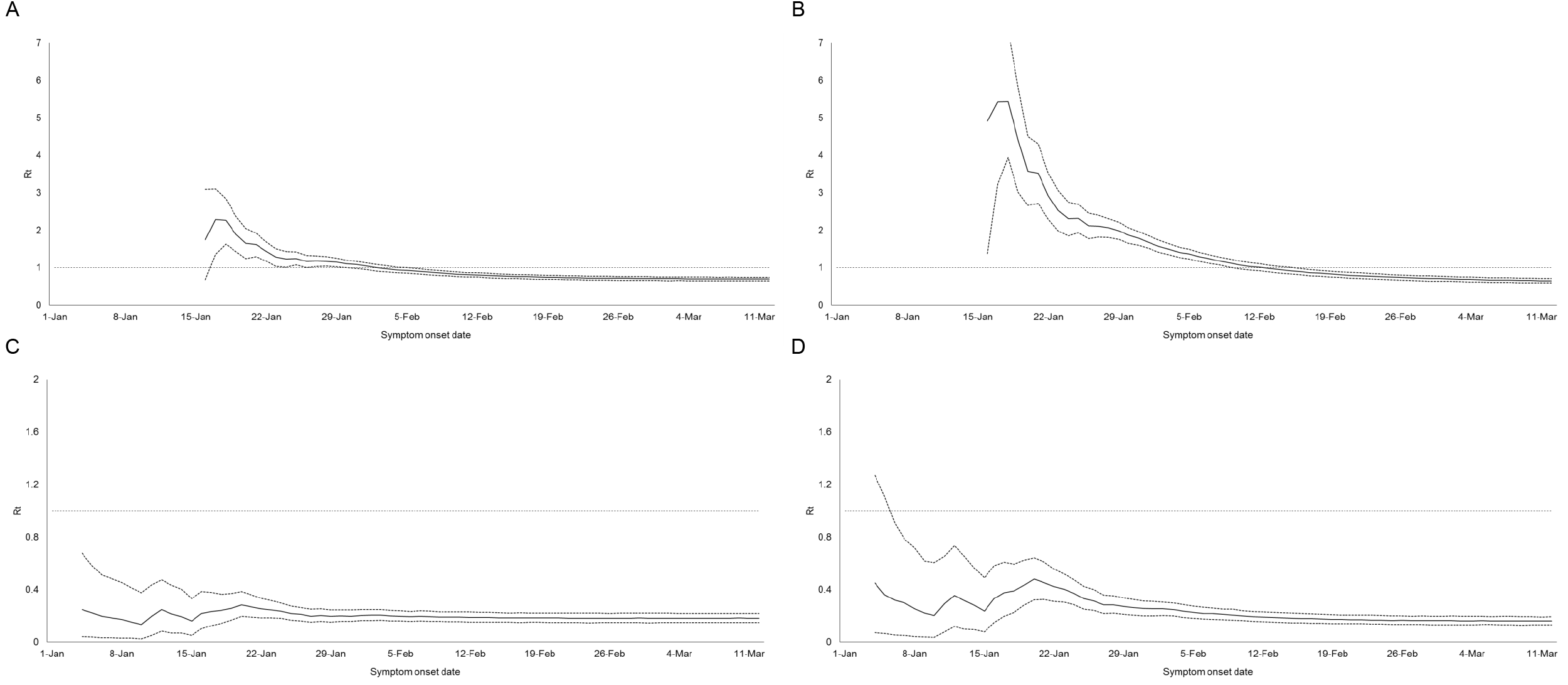
Time-varying reproduction number (*R*_*t*_) assuming a shorter (A, C) and longer serial interval (B, D) in Hangzhou (A, B) and Shenzhen (C, D) respectively.

## Discussion

Monitoring the transmission dynamics as well as the *R*_*t*_ of the disease is useful in 1. determining whether there is sustained community transmission in a population and 2. evaluating whether the control measures taken are adequate to control the local transmission at a specific time [26-28]. In this study, we estimated the imported-cases-adjusted *R*_*t*_ of COVID-19 in the first wave of epidemic in Hangzhou and Shenzhen, two Chinese cities with different transmission dynamics of imported cases. According to our results, the disease transmissibility in both cities gradually decreased over time especially after the lockdown of Wuhan. Credits to the community lockdown, the transmission was interrupted within a short period of time in Hangzhou. In line with Lai et al. [29], inter-city travel restrictions and other social distancing interventions could slow down the epidemics outside Wuhan.

We showed that local transmission of COVID-19 seeded by imported cases was not sustained in Shenzhen and we speculated it was likely due to the decrease in number of local susceptibles during Chunyun. Of the 13 million residents in Shenzhen, around 65% of its population were migrants which ranked top in China, and most of the residents have returned to their home towns starting from 10 January for celebration of Lunar New Year [17,30]. Given the number of local susceptibles decreased as a large number of residents left Shenzhen during Chunyun, fewer transmission chains could be established during this period even though Shenzhen is highly connected to Wuhan for most of the times. Tian et al. [11] indicated that Chinese cities that implemented control measures before officials confirmed the first case were more likely to have fewer cases. Even though the first case in Shenzhen was confirmed on 19 January (illness onset on 3 January), we believe Shenzhen is an exception owing to its special population characteristics. In line with Kucharski et al. [31], the case study of Shenzhen supported that introducing one to several cases to a new city may not necessarily lead to an outbreak.

Comparing with Shenzhen, Hangzhou is not just a highly populated city with fewer migrants, but is also geographically closer to Wuhan. By implementing strict control measures before cases emerged, local transmissions were comparatively easier to be seeded in Hangzhou. Compared to the transmission dynamics in Wuhan [31], *R*_*t*_ in Hangzhou displayed a consistently declining trend staring from mid-January. This may be due to the increased awareness on the use of personal protective measures against COVID-19 after noticing the unknown pneumonia outbreak in Wuhan in early January through social media. Similar transmission dynamics was also reported in Shaanxi province [32].

In this study, we showed that even though more cases were reported in Shenzhen when compared to Hangzhou, they exhibited different transmission dynamics of COVID-19 when the risk of imported cases was accounted in the estimation. With more local cases emerged during the initial phase of the epidemic, the disease sustained in Hangzhou before the lockdown of Wuhan and the initiation of outbreak response. For cities or provinces with a large proportion of imported cases, failing to differentiate them from local cases during *R*_*t*_ calculation may lead to overestimation of transmissibility in a local population [33], and thus, affect the planning of mitigation measures. The importance of accounting for imported cases has been demonstrated in another study that looked into Middle East Respiratory Syndrome in Saudi Arabia [21].

Our study has several major limitations. Recent studies have successively reported that some patients infected with COVID-19 might infect others before their symptom onset [24,34]. Disease transmission during the pre-symptomatic stage implies the possibility of having a negative value of serial interval. This would affect the formulation of estimation equation since the distributional assumption did not gave it a corresponding probability. Alternatively, a generation interval could be used but it is usually hard to be estimated due to unobserved infectiousness onset. Nevertheless, our sensitivity analysis demonstrated that the variation of serial interval within a reasonable range did not affect our main conclusion. Another limitation is the underreporting of confirmed cases due to the unavailability of virological testing in the early stage of epidemic. According to an early investigation [35], an estimate of 75_thousand individuals were found to be infected in Wuhan as of 25 January and we believe similar underreporting was likely to occur in our setting. With detailed data available on reporting rates and serological surveillance, our analytic frame can be extended to a more complex context that incorporates these factors. In addition, our estimates shall be refined if more updated knowledge of the pathogen is available.

In conclusion, we showed the lockdown measures and local outbreak responses helped reduce the potential of local transmission in Hangzhou and Shenzhen. The low transmission intensity of COVID-19 in Shenzhen in early January was likely due to the decrease in number of local susceptibles as residents left the city for Chunyun. We also highlighted that cities with similar epidemic trend could have different transmission dynamics given the variation in imported cases.

## Data Availability

All data were downloaded from the official websites and they were publicly available.

## Acknowledgments

We thank the physicians and staffs at Hangzhou, Huzhou, Jiaxing Wenzhou, Shaoxing, Ningbo, Quzhou, Jinhua, Zhoushan, Lishui, Taizhou Municipal Center for Disease Control and Prevention for their support and assistance with this investigation.

## Ethics

All data were collected via official websites and publicly available so neither ethical approval nor individual consent was not applicable.

